# PD-1/PD-L1 immune checkpoint therapy demonstrates favorable safety profile in patients with autoimmune liver disease

**DOI:** 10.1101/2023.07.17.23292183

**Authors:** Lorenz Kocheise, Ignazio Piseddu, Joscha Vonderlin, Eric T Tjwa, Gustav Buescher, Lucy Meunier, Pia Goeggelmann, Francesca Fianchi, Jérôme Dumortier, Mar Riveiro-Barciela, Tom JG Gevers, Benedetta Terziroli Beretta-Piccoli, Maria-Carlota Londoño, Sona Frankova, Thomas Roesner, Vincent Joerg, Constantin Schmidt, Fabian Glaser, Jan P Sutter, Thorben W Fründt, Ansgar W Lohse, Samuel Huber, Johann von Felden, Marcial Sebode, Kornelius Schulze

**Author notes:** **Corresponding Author:** Lorenz Kocheise, I. Department Medicine, University Medical Center Hamburg-Eppendorf, Martinistr. 52, Hamburg 20246, Germany. These authors contributed equally to this work (Marcial Sebode, Kornelius Schulze).

## Abstract

**Background:** Immune checkpoint inhibitors (ICI) have revolutionized the treatment of many malignancies in recent years. However, immune-related adverse events (irAE) are a frequent concern in clinical practice. The safety profile of ICI for the treatment of malignancies in patients diagnosed with autoimmune liver disease (AILD) remains unclear. Due to this uncertainty, these patients were excluded from ICI clinical trials and ICI are withheld from this patient group. In this retrospective multicenter study, we assessed the safety of ICI in patients with AILD.

**Methods:** We contacted tertiary referral hospitals for the identification of AILD patients under ICI treatment in Europe via the European Reference Network on Hepatological Diseases (ERN RARE-LIVER). Fourteen centers contributed data on AILD patients with malignancies being treated with ICI, another three centers did not treat these patients with ICI due to fear of irAEs.

**Results:** In this study, 22 AILD patients under ICI treatment could be identified. Among these patients, 12 had primary biliary cholangitis (PBC), five had primary sclerosing cholangitis (PSC), four had autoimmune hepatitis (AIH), and one patient had an AIH-PSC variant syndrome. Eleven patients had hepatobiliary cancers and the other 11 patients presented with non-hepatic tumors. The applied ICIs were atezolizumab (n=7), durvalumab (n=5), pembrolizumab (n=4), nivolumab (n=4), spartalizumab (n=1), and in one case combined immunotherapy with nivolumab plus ipilimumab. Among eight patients who presented with grade 1 or 2 irAEs, three demonstrated liver irAEs. Cases with grades ≥ 3 irAEs were not reported. No significant changes in liver tests were observed during the first year after the start of ICI.

**Conclusions:** This European multicenter study demonstrates that PD-1/PD-L1 inhibitors appear to be safe in patients with AILD. Further studies on the safety of more potent dual immune checkpoint therapy are needed. We conclude that immunotherapy should not categorically be withheld from patients with AILD.

## Introduction

Immune checkpoint inhibitors (ICI) have significantly improved the prognosis of patients with advanced tumor stages in recent years^1-6^. Patients with concomitant autoimmune liver diseases (AILD) were excluded from these pivotal clinical trials due to the concerns about immune-related adverse events (irAEs). However, patients with AILD, including autoimmune hepatitis (AIH), primary biliary cholangitis (PBC) and primary sclerosing cholangitis (PSC), have a significantly increased risk of developing hepatocellular carcinoma (HCC) at the stage of liver cirrhosis or biliary tract cancer (BTC) as in the case of PSC^7-9^. While ICI therapy is now the approved first-line treatment for these hepatobiliary malignancies, it is rarely offered to patients with AILD due to safety concerns and a lack of clinical data^4-6^.

Checkpoint inhibitor-induced liver injury (CHILI) is a common manifestation of irAE and occurs in 5-10% of patients on PD-1/PD-L1 monotherapy and in up to 30% of all patients under anti-CTLA-4 combination therapy^10,11^. For patients with HCC, increased levels of aspartate-aminotransferase (AST) were detected in about 20% of all patients under PD-L1 therapy^4^. Several cases of severe CHILI, including death due to acute liver failure, have been reported^12,13^. Different forms of CHILI have been identified including immune-related hepatitis (irHepatitis) and immune-related sclerosing cholangitis (irSC), mimicking the entire spectrum of autoimmune diseases of the liver^14-17^. It is unclear whether individuals with AILD are at an increased risk for the development of these hepatic irAE, given the potential amplification by autoimmune susceptibility.

While retrospective observational data on ICI treatment are now available for other autoimmune diseases, data for patients with AILD is still lacking^18,19^. This uncertainty has led to reluctance of hepatologists and oncologists to consider ICI therapy for these patients. However, the results of a recent study supports that CHILI is histologically and clinically distinct from AILD and thereby probably does not share the same pathophysiological mechanisms^20^. This suggests that AILD patients under ICI treatment may not be at increased risk for irAE. In this multinational, multicenter retrospective study, we evaluated the clinical safety and response profile of ICI therapy in tumor patients with concomitant AILD.

## Patients and Methods

### Patients

We contacted tertiary referral hospitals for AILD in Europe via the European Reference Network on Hepatological Diseases (ERN RARE-LIVER). Fourteen centers treated malignancies with ICI in patients with AILD, three other centers stated that they decided not to treat AILD patients with ICI due to the fear of irAEs, and 13 other centers were not able not identify any AILD adult patients being treated with ICI at the respective center. All AILD patients who received at least one dose of an ICI for the treatment of hepatic or extrahepatic malignancy were included. Anonymized data was collected. In accordance with the local ethics committee (#2023-300290-WF), no ethical approval was necessary for this anonymous online survey. The clinical trial was conducted in accordance with Good Clinical Practice (GCP) guidelines and the Declaration of Helsinki.

### Study design

Baseline patient demographics and clinical characteristics, including gender, age at the start of immunotherapy, AILD type, cancer type, and laboratory and radiological response data were assessed with an online survey tool provided by the EU for ERNs (https://ec.europa.eu/eusurvey). The severity of irAE was graded according to the current Common Terminology Criteria for Adverse Events (CTCAE). Inclusion criteria were the diagnosis of an AILD with at least one dose of ICI treatment. No reported patients were excluded from this study. Alternative causes of liver injury in case of irAE were excluded by the local clinicians of the corresponding tertiary referral center.

### Statistical analysis

Paired t-test was used to compare laboratory values before and after treatment. Duration of ongoing ICI therapy and overall survival (OS) were analyzed using the log-rank test and plotted with Kaplan-Meier curves. For all statistical analyses, p-values below 0.05 were considered significant. All statistical analyses were conducted on GraphPad Prism (Version 9.5.1).

## Results

### Patient characteristics

22 AILD patients with malignancies being treated with ICI were identified. Baseline characteristics are presented in **Table 1**. Of the 22 patients, six (27.3%) were male and 16 (72.7%) were female. The mean age at the start of immunotherapy was 63.4 years. Twelve patients (54.4%) had PBC, five (22.7%) had PSC, four (18.2%) had AIH, and one (4.5%) patient had an AIH-PSC variant syndrome. Five (19%) patients had a history of other autoimmune diseases including autoimmune thyroiditis, autoimmune hemolytic anemia, Sjogren’s syndrome, ulcerative colitis, and rheumatoid arthritis. Eight (36.4%) patients had underlying liver cirrhosis. Eleven (50.0%) patients had hepatobiliary cancers and the remaining 11 patients presented with extrahepatic tumors. The latter group included two (9.1%) patients with liver metastases. The most frequent tumor types were HCC (n=6), BTC (n=5), non-small cell lung cancer (n=4) and malignant melanoma (n=2). Other tumor entities were breast cancer (n=1), esophageal cancer (n=1), small cell lung cancer (n=1), renal cell cancer (n=1), and head and neck cancer (n=1). The applied ICIs were atezolizumab (n=7), durvalumab (n=4), pembrolizumab (n=4), nivolumab (n=4), spartalizumab (n=1), and in one case dual ICI therapy with nivolumab plus ipilimumab (CTLA-4 inhibitor).

**Table 1.** Demographic characteristics of AILD patients.

| Baseline Characteristics | Patients, n = 22 (%) |
| --- | --- |
| **Gender** |  |
| Male | 6 (27.3) |
| Female | 16 (72.7) |
| **Age** |  |
| Mean ± SD | 63.4 ± 13.3 |
| **Liver cirrhosis** |  |
| Yes | 8 (36.4) |
| No | 14 (63.6) |
| **Autoimmune liver disease** |  |
| Autoimmune hepatitis | 4 (18.2) |
| Primary biliary cholangitis | 12 (54.5) |
| Primary sclerosing cholangitis | 5 (22.7) |
| Variant syndrome AIH-PSC | 1 (4.5) |
| **Other autoimmune disease** |  |
| Autoimmune hemolytic anemia | 1 (4.5) |
| Autoimmune thyroiditis | 1 (4.5) |
| Rheumatoid arthritis | 1 (4.5) |
| Sjögren syndrome | 1 (4.5) |
| Ulcerative colitis | 2 (9.1) |
| None | 17 (77.3) |
| **Tumor type** |  |
| Hepatobiliary cancer | 11 (50.0) |
| Non hepatobiliary cancer | 11 (50.0) |
| Liver metastases present | 2 (9.1) |
| No liver metastases present | 9 (40.9) |

The following tumor diseases were registered: Hepatocellular cancer (n=6), biliary tract cancer (n=5), non-small cell lung cancer (n=4), malignant melanoma (n=2), breast cancer (n=1), esophageal cancer (n=1), small cell lung cancer (n=1), renal cell cancer (n=1), and head and neck cancer (n=1).

### Baseline immunosuppression and duration of immune checkpoint inhibitor treatment

At the time of ICI initiation, nine (40.9% of all patients; 6/12 of PBC, and 3/5 PSC patients) patients were treated with ursodeoxycholic acid (UDCA) and one PBC patient was treated with obeticholic acid (OCA). Six patients (27.3% of all patients; 3/4 AIH patients, 2/12 PBC patients, and one patient with AIH-PSC variant syndrome) received systemic immunosuppressive therapy. Seven (31.8% of all patients; 1/4 AIH patient, 4/12 PBC patients, and 2/5 PSC patients) patients did not receive any therapy for their AILD at the time of initiation of ICI therapy. Details on the treatment of AILD patients before and during ICI are depicted in **Table 2**. The timelines of ICI treatment regimens are illustrated in **Figure 1**. The median duration of ongoing ICI therapy was 35 weeks (range: 3 – 61 weeks), and the median overall survival for all patient was 76 weeks. The influence of immunosuppressive medication on the duration and sustainability of ICI therapy was investigated using Kaplan-Meier curves. Patients treated with immunosuppressive therapy, UDCA or OCA and patients without treatment of AILD were grouped and the corresponding Kaplan-Meier curves are plotted in **Figure 2**. There was no significant difference between the groups regarding time on ICI therapy.

**Table 2.** Immune-related adverse events and immunosuppression.

| Baseline characteristics | Patients, n = 22 (%) |
| --- | --- |
| **irAE** |  |
| Grade 1 | 3 (13.6) |
| Grade 2 | 5 (22.7) |
| None | 14 (63.6) |
| **Liver irAE** |  |
| Grade 1 | 2 (9.1) |
| Grade 2 | 1 (4.5) |
| **Non-Liver irAE** |  |
| Colitis | 1 (4.5) |
| Pneumonitis | 1 (4.5) |
| Inflammatory arthritis | 1 (4.5) |
| Rash | 2 (9.1) |
| **Treatment regimen before ICI<sup>#</sup>** |  |
| Mycophenolate mofetil | 1 (4.5) |
| Azathioprine | 4 (18.1) |
| Corticosteroids | 2 (9.1) |
| Methotrexat* | 1 (4.5) |
| UDCA | 9 (40.9) |
| Obeticholic acid | 1 (4.5) |
| No treatment for AILD | 7 (31.8) |
| **Treatment adjustment due to pre-existing autoimmune disease or irAE<sup>#</sup>** |  |
| Start Hydroxychloroquine* | 1 (4.5) |
| Start or increase of corticosteroids | 4 (18.1) |
| Start UDCA | 1 (4.5) |
| Change from obeticholic acid to Fenofibrate | 1 (4.5) |
| Stop Azathioprine | 1 (4.5) |
| No change in treatment | 16 (72.7) |
| **Monoclonal antibody used** |  |
| Atezolizumab | 7 (33.3) |
| Durvalumab | 5 (19.0) |
| Pembrolizumab | 4 (19.0) |
| Nivolumab | 4 (19.0) |
| Nivolumab + Ipilimumab | 1 (4.8) |
| Spartalizumab | 1 (4.8) |

#Multiple treatments possible \*Treatment for Rheumatoid arthritis

**Figure 1.**
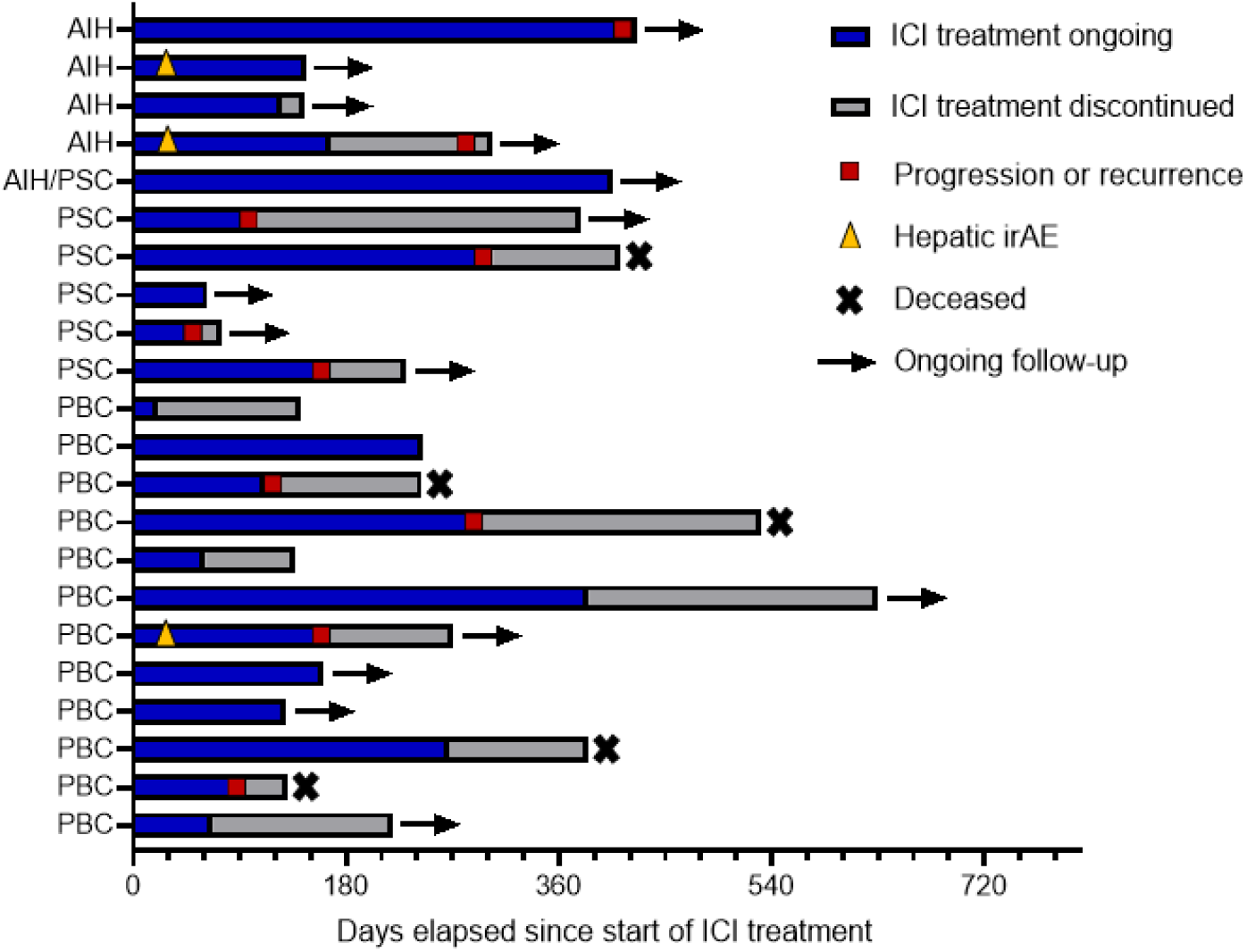
Therapy timeline of individual patients. ICI, Immune checkpoint inhibitor treatment; AIH, Autoimmune hepatitis; PSC, primary sclerosing cholangitis; PBC, primary biliary cholangitis. Hepatic irAEs of grades I and II occurred. A flare of the underlying autoimmune disease could not be distinguished from an irAE.

**Figure 2.**
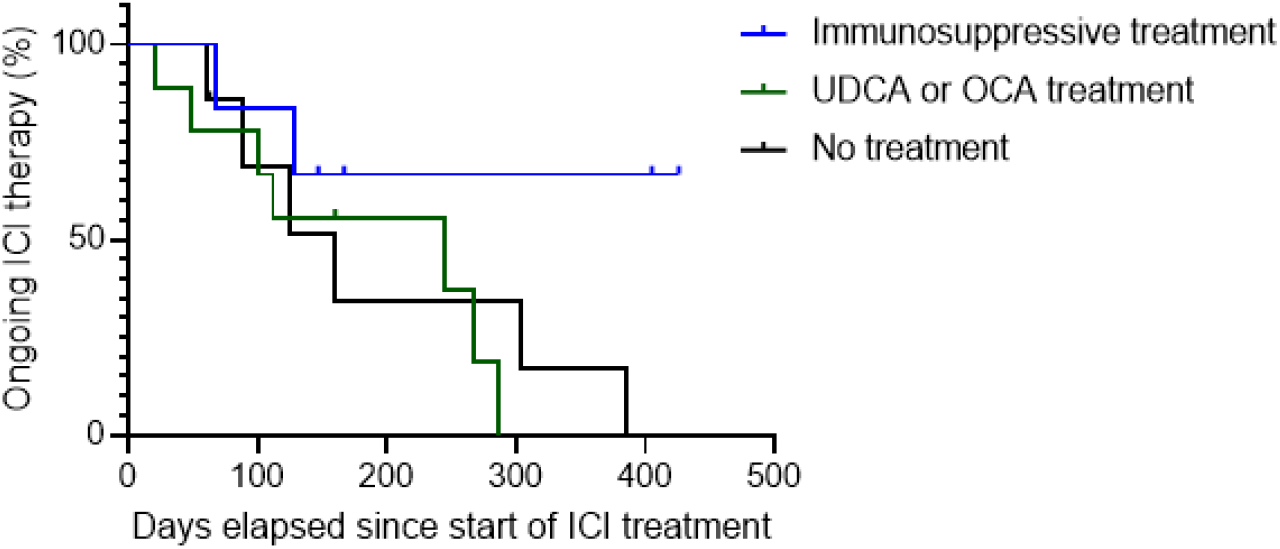
Kaplan-Meier curves for time on immune checkpoint inhibitor treatment (ICI). There was no significant differences using the log-rank test.

### Immune-related adverse events and changes in immunosuppressive treatment

IrAEs and immunosuppression before and during ICI treatment are depicted in **Table 2**. Among the analyzed patients, eight (36.4%) developed irAEs during ICI therapy. Three of these were grade 1 irAEs, while five were grade 2 irAEs. Out of the total eight irAEs observed, three specifically affected the liver. These hepatic irAEs consisted of two cases classified as grade 1 and one case classified as grade 2. All hepatic irAEs occurred in the first two months after ICI initiation and did not lead to discontinuation of ICI therapy.

Of these three cases with increased liver enzymes under ICI, one patient was diagnosed with PBC after the beginning of ICI therapy, although the diagnosis of PBC was already indicated by increased ALP levels before ICI treatment initiation. The diagnosis of PBC was then further supported by the detection of antimitochondrial autoantibodies and elevated IgM levels. The patient did not receive any treatment for AILD prior to starting ICI therapy. Liver enzyme elevations, indicative of an irAE, manifested after 2 weeks and ALT and AST levels peaked at 100 and 145 IU/l respectively. After initiating therapy with UDCA, the patient’s liver enzymes normalized within a few weeks. ICI therapy was continued until tumor progression. However, since the ALT and AST elevation could not be distinguished from an irAE, the incident was considered as grade 2 irAE.

The other two cases with grade 1 liver irAE occurred in AIH patients. One patient has been under treatment with mycophenolate mofetil (1g bid) for AIH for several years before ICI therapy, ALT and AST levels were normal before ICI initiation, while IgG levels were not known. Liver enzyme elevations, indicative of an irAE, manifested after four weeks and ALT levels peaked at 80 IU/l. After initiation of an oral prednisolone therapy with 15 mg per day, elevated liver enzymes after ICI treatment normalized within four months, and ICI therapy was continued until the completion of the adjuvant treatment line without another increase of liver enzymes. The other patient received azathioprine (100 mg per day) and prednisone (5 mg per day) for 10 months before the initiation of ICI therapy. Liver enzyme elevations, indicative of an irAE, manifested after three weeks and ALT and AST levels both peaked at 58 IU/l. After an increase of the prednisone dosage to 10 mg per day liver enzymes normalized within three weeks. Prednisone dosage was reduced to 7.5 mg per day and ICI therapy is still ongoing after half a year without another hepatic irAE. Furthermore, it remains unclear whether the increase of liver enzymes in patients with AIH represented a flare of AIH or a mild episode of CHILI. Details of all AIH patients are presented in **Table 3**. While all AIH patients had normal ALT levels at the beginning of ICI therapy, IgG levels ranged from 5.3 – 30.8 g/l. 75% of all AIH patients received immunosuppressive treatment before the start of ICI therapy. Notably, the AIH patient who did not receive any immunosuppressive treatment had the highest IgG levels among all AIH patients and did not experience any irAEs during ICI therapy. In most patients (72.7%), treatment for AILD was not changed during ICI therapy. In one patient with concomitant rheumatoid arthritis, immunosuppressive therapy was switched from methotrexate to hydroxychloroquine, due to inflammatory arthritis. Two patients with non-hepatic irAE received a short-term steroid treatment until symptoms resolved. In one patient with AIH, maintenance therapy with azathioprine (50 mg per day) was discontinued during ICI treatment and no increase of liver enzymes was detected during the observational period of 14 months.

**Table 3:** Characteristics of patients with autoimmune hepatitis.

| Baseline characteristics | Patients, n = 4 (%) |
| --- | --- |
| **Treatment regime before ICI<sup>#</sup>** |  |
| Corticosteroids (5 mg per day) | 1 (25) |
| Mycophenolate mofetil (2 g per day) | 1 (25) |
| Azathioprine* | 2 (50) |
| None | 1 (25) |
| **IgG levels before ICI treatment** |  |
| Median (Range) in g/l | 10.1 (5.3 - 30.8) |
| Not available | 1 (25) |
| **ALT levels before ICI treatment** |  |
| Median (Range) in IU/l | 26.3 (18.0 - 33.0) |
| **Time on immunosuppressive treatment for AIH before ICI therapy** |  |
| Mean (Months) ± SD | 30.6 ± 6.0 |
| **Treatment adjustment during ICI therapy** |  |
| Start of corticosteroids (15 mg per day) | 1 (25) |
| Increase of corticosteroids (5 mg per day) | 1 (25) |
| Stop of immunosuppression | 1 (25) |
| No treatment adjustment | 1 (25) |
| **Highest ALT levels during ICI treatment** |  |
| Mean (Range) in IU/l | 54.75 (40 – 80) |
| **Weeks until normalization of liver enzymes after irAE** |  |
| Mean (Range) | 10 (2 – 18) |

<sup>#</sup>Multiple treatments possible, \*Azathioprine: 100 mg and 50 mg per day respectively.

### Liver function during immune checkpoint inhibitor treatment

Liver function parameters were compared to exclude a progression of autoimmune liver disease and subsequent impaired liver function after ICI therapy **(Figure 3)**. The laboratory values INR, total bilirubin, ALP, and ALT levels were compared before and after the start of ICI therapy and after 1, 3, 6 and 12 months of treatment, although the availability of complete datasets decreased over time. There was no significant difference between the different time points. A subgroup analysis of AIH, PSC, and PBC patients also showed no significant difference during the treatment timeline.

**Figure 3.**
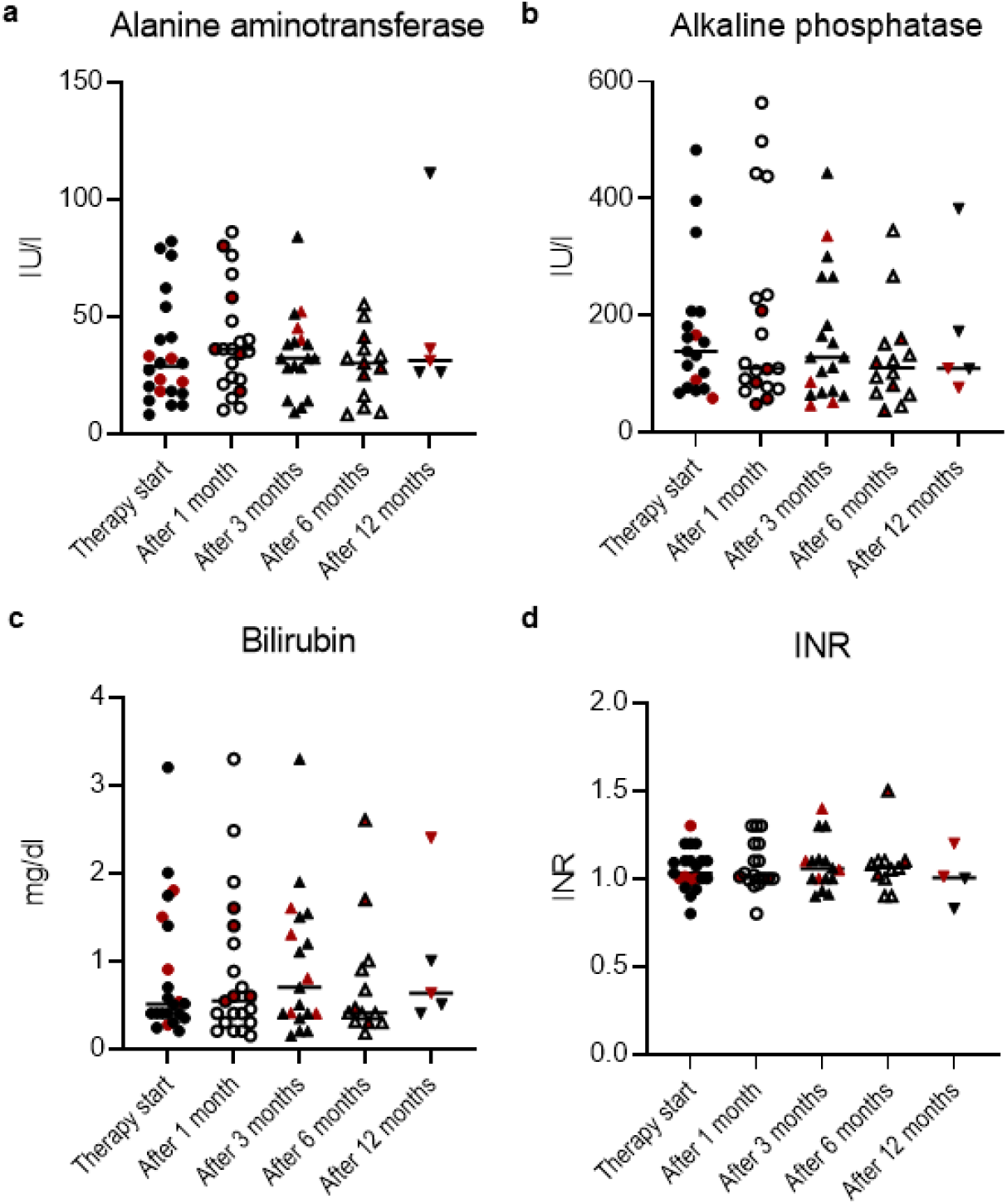
ALT, ALP, total bilirubin levels, and INR during immune checkpoint inhibitor treatment. Laboratory values of AIH and AIH/PSC patients were highlighted in red. There was no significant difference at any time point. Values before therapy initiation were compared with individual values during therapy using paired t-tests. A subgroup analysis of AIH, PSC, and PBC patients also showed no significant difference during the treatment timeline.

## Discussion

While retrospective multicenter data for the course of extrahepatic autoimmune diseases under ICI therapy have already been published, such data is very limited for the treatment of malignancies in AILD^19,21-24^. Only very few single patients with AILD were analyzed in these studies. In a retrospective multicenter study including 85 patients with different pre-existing autoimmune diseases being treated with PD-1/PD-L1 inhibitors, one patient with PSC was included^25^. Another multicenter study of 112 patients with a wide variety of autoimmune diseases reported one case with AIH and two cases with PBC. One of the patients with PBC developed acute liver failure, but its etiology remains unclear^21^. Despite an increased rate of autoimmune flares, ICI therapy has demonstrated reasonable safety in many autoimmune diseases. Retrospective studies focusing on extrahepatic irAE have been published for rheumatoid arthritis, psoriasis, or inflammatory bowel disease^27-29^. However, the heterogeneity of autoimmune diseases included in these retrospective studies is a relevant limitation^26^.

Although the respective immunological mechanisms remain elusive for both disease groups, AILD and ICI-induced liver injury show some similarities, but also several distinct features. Different manifestations of hepatic irAEs are grouped under the term CHILI: hepatitic, cholestatic or mixed pattern have been proposed^14,15^. In addition to the more frequent hepatic involvement, sclerosing cholangiopathies (irSC) have also been detected, with steroid response rates as low as about 10%^13,17,30^. The appearance and non-responsiveness to steroids represent similarities between PSC and irSC. In contrast to this, histological studies suggest that hepatitic CHILI and AIH are more distinct entities^20^, while classical histological features of AIH include interface hepatitis and plasma cell infiltration, liver injury due to ICI is characterized by granulomatous hepatitis including endotheliitis or lobular hepatitis^31^. Therefore, it remains unclear whether AILD and hepatic irAEs share immunological mechanisms and consequently, whether pre-existing AILD represents a risk factor for more frequent or more severe hepatic irAEs.

In this multicenter retrospective study, we analyzed the safety profile of ICI in AILD patients. Most importantly, no severe forms of CHILI in terms of irAE grade ≥ 3 occurred. Overall, only three patients showed laboratory values that could indicate CHILI. In two out of three cases, a mild flare of AIH could not be distinguished from irAE grade 1, whereas the third case was a patient with untreated PBC. None of these patients required interruption of ICI therapy, and liver enzyme levels normalized rapidly in response to immunomodulatory treatment. For the study cohort, we could not detect a significant increase in mean liver enzyme levels after initiation of checkpoint therapy. In addition, liver function parameters such as INR or bilirubin did not change significantly during ICI therapy, which is of particular importance for those 35% of all patients being included at the stage of liver cirrhosis. These patients may be at particular risk of developing acute on chronic liver failure due to CHILI. During the observation period, we did not detect an increase in cholestasis parameters in patients with PSC. However, it is important to note that the observation period was likely too short to evaluate the potential progression of bile duct sclerosis in most patients.

The limitations of this study are primarily due to its retrospective design. Given the limited patient numbers, a comprehensive investigation of further potential factors influencing the occurrence of hepatic irAE, such as cirrhosis stage, tumor type, type of ICI inhibitor, and pre-existing immunosuppressive therapy, could not be conclusively analyzed. Another limitation of this study is that only one patient received combined immunotherapy with CTLA-4 and PD-1/PD-L1 blockade. Dual therapy with PD-1/PDL1 and CTLA-4 inhibitors has been associated with a significantly higher risk of irHepatitis when compared to monotherapy^2,3^. Further data on the safety of dual ICI therapy in patients with AILD is urgently needed, especially when the recent approval of combined immunotherapy with the CTLA-4 inhibitor tremelimumab and the PD-L1 inhibitor durvalumab for HCC is being considered. In addition, no patient was treated with the FDA and EMA approved LAG-3 immune checkpoint inhibitor relatlimab and the data on the safety of this immune checkpoint for patients with autoimmune diseases is missing^32^. Nevertheless, ICI monotherapy with PD-1/PD-L1 inhibitors is the first-line therapy for most immune-oncology treatment indications. In addition, previous studies suggest that the risk of irSC is primarily attributed to PD-1/PD-L1 inhibitors^33^. Although ICI therapy was generally well tolerated in this study and hepatic irAEs did not result in the discontinuation of ICI therapy, it is important to note that two out of four AIH patients experienced a transient increase in transaminase levels characterized as irAE grade I. In both cases, an irAE could not be distinguished from an AIH flare and ICI therapy was not discontinued due to the irAE. Flare of pre-existing autoimmune disease after initiation of ICI therapy have been described previously for other entities. However, additional data is needed for AIH. Meanwhile, ICI therapy should not be discarded from AIH patients, although close monitoring is still required. The impact of ICI therapy on the chronic progression of underlying AILD remains uncertain. This study observed no impairment of liver function tests within the first year following the initiation of ICI therapy. However, it should be noted that liver elastography, which could provide insights into increasing fibrosis of the liver, was rarely performed in this patient cohort due to the advanced tumor stages being treated. Therefore, the possibility of progressive liver fibrosis cannot be entirely excluded in this context. Since adjuvant and neoadjuvant ICI therapy with curative intent is becoming more and more widespread, the long-term effect of ICI therapy for AILD needs to be investigated in the future. The strength of this study lies in its multicenter multinational approach, which enables the collection of a diverse range of cases of AILD under ICI treatment. In contrast to previous studies, this study focuses on immune-mediated autoimmune diseases of one organ and did not include the wide range of multisystemic autoimmune diseases.

In conclusion, this retrospective multicenter suggests that ICI treatment of malignancies seems to be safe in patients with autoimmune liver diseases. ICI treatment should not be categorically withheld from these patients, but close monitoring is still required. Future studies including more patients, particularly those with autoimmune hepatitis and dual ICI therapy, are needed to further analyze the risk profile of ICI treatment in these patients.

## Contributions

Study concept and design: LK, SH, AWL, JvF, MS, KS

Acquisition of data: LK, IP, JV, ETT, GB, LM, PG, FF, JD, MRB, TJGG, BTBP, MCL, SF, TR, VJ, CS, FG, JPS, TWF, MS and KS

Analysis of data: LK, MS, KS

Drafting of the manuscript: LK, AWL, MS, KS

All authors approved the final version of the manuscript after critical revision.

## Ethics declarations

In accordance with the local ethics committee (#2023-300290-WF), no ethical approval was necessary for this anonymous online survey. The clinical trial was conducted in accordance with Good Clinical Practice (GCP) guidelines and the Declaration of Helsinki. All data was gathered by the treating hospital and subjected to anonymous analysis.

## Data availability

All data relevant to the study are included in the article.

